# Late-Onset Neutropenia in a Single-Center, Retrospective Cohort of Central Nervous System Autoimmunity Patients Treated with Anti-CD20

**DOI:** 10.64898/2026.08.14.26360444

**Authors:** Ahmed H. Althobaiti, Nora Abanmi

**Affiliations:** Neurology Department, King Saud Medical City, Riyadh First Health Cluster, Riyadh, Saudi Arabia, 12746

**Keywords:** late-onset neutropenia, multiple sclerosis, NMOSD, rituximab, ocrelizumab, CNS autoimmunity

## Abstract

**Background:** Late-onset neutropenia (LON) is an infrequently reported, unpredictable side effect of anti-CD20 therapy, with incidence varying by agent, diagnosis, and screening protocol.

**Objective:** The primary objective of this cross-sectional, retrospective study was to estimate the proportion of patients who developed LON over 13 months (April 2023–April 2024).

**Methods:** Consecutive adult patients diagnosed with central nervous system (CNS) autoimmunity who received at least one rituximab(RTX) or ocrelizumab(OCR) infusion between January 2016 and March 2024 were included; patients who switched to another immunotherapy, had no post-treatment blood draw, or had unverifiable infusion records were excluded. LON events were assessed using all post-treatment CBCD blood draws during this period.

**Results:** A total of 171 patients were enrolled: 141 received rituximab and 30 received ocrelizumab. A total of 319 post-treatment blood tests were performed. Sixteen patients (16/171) had neutropenia (9.4%, 95% CI 5.8–14.7): 12 on rituximab (8.5%) and 4 on ocrelizumab (13.3%; p=0.487). LON occurred at a median of 158 days (130–188) since the last infusion. All patients were asymptomatic, mostly had Grade 1 neutropenia (15/16, 93.8%). BMI (22.2 vs. 27.5 kg/mš, p=0.001) and prior natalizumab exposure (37.5% vs. 14.2%, p=0.023) were significantly different between neutropenic and non-neutropenic patients.

**Conclusion:** The proportion of patients with LON in this cohort was higher than most previously reported, with all cases asymptomatic. Lower BMI and prior natalizumab exposure emerged as potential risk factors warranting further investigation. Larger, prospective studies with standardized surveillance are needed to establish the true frequency and risk factors.

## 1 Introduction

Anti-CD20 monoclonal antibodies are widely used to treat central nervous system (CNS) autoimmune disorders (Stathopoulos, and Dalakas 2022). One of their infrequently reported and unpredictable adverse effects is late-onset neutropenia (LON) (Baker et al. 2024).

LON was first described in the context of rituximab therapy in 2003 (Chaiwatanatorn et al. 2003), and is operationally defined as an absolute neutrophil count (ANC) 1.5 Œ 10^9^/L occurring more than 4 weeks after the last infusion, without an alternative explanation, graded 1–4 by severity adapted from the Common Terminology Criteria for Adverse Events (CTCAE) version 6.0.

Reported LON incidence varies by anti-CD20 agent, underlying diagnosis, testing frequency, and treatment duration and intensity, with lower rates generally reported in autoimmune disease compared to hematologic malignancy (Wolach et al. 2012; Salmon et al. 2015). Multiple case reports describe LON following anti-CD20 therapy for CNS autoimmunity (Plate et al. 2014; Cohen 2019; Zanetta et al. 2020; Baird-Gunning et al. 2021; Maniscalco et al. 2021; Marrodan et al. 2021; Rauniyar et al. 2022).

Ideally, rigorous and systematic evaluation of large cohorts of CNS-autoimmunity patients treated with anti-CD20 would yield a more robust estimate of LON occurrence.

Rigal et al. (Rigal et al. 2022) prospectively assessed ANC every 2 weeks over 6 months in a cohort of multiple sclerosis, NMOSD, and MOGAD patients, applying a stricter threshold (ANC 1.0 Œ 10^9^/L, equivalent to Grade 2 or worse) and identified 2/152 cases; however, a 6-month window presumes LON is predominantly an early phenomenon and may miss later presentations.

Waldrop et al. (Waldrop et al. 2025) reported a lower incidence rate of 0.62 (95% CI 0.45–0.85) per 100 person-years over a much longer period (January 6, 2006–November 2, 2015) using an EHR-based query; however, such a design cannot ensure complete case capture if patients sought care outside the reporting system, and is likely biased toward symptomatic neutropenia given its unreported screening frequency.

Together, these studies leave a persistent gap: few evaluations have combined adequate follow-up duration, complete case ascertainment, and transparent reporting of testing frequency.

The primary objective of this retrospective, cross-sectional observational study was to estimate the proportion of patients who developed LON in our cohort of CNS autoimmunity over a 13-month period (April 2023–April 2024), as an exploratory analysis to motivate more extensive evaluation of this complication.

## 2 Methods

This was a single-center, retrospective, cross-sectional observational study of patients with CNS autoimmunity treated with rituximab or ocrelizumab and evaluated for LON over a 13-month period. Consecutive sampling was used to enroll all adult patients with CNS autoimmunity diagnosed according to standard criteria (multiple sclerosis: McDonald 2017 (Thompson et al. 2018); Neuromyelitis Optica Spectrum Disorders(NMOSD): International Panel for NMO Diagnosis 2015 (Wingerchuk et al. 2015) ; Susac syndrome: Kleffner et al. 2016 (Kleffner et al. 2016) ; Autoimmune Encephalitis (AE): Graus et al. 2016 (Graus et al. 2016) ;Myelin Oligodendrocyte Glycoprotein Associated Disease (MOGAD): 2023 International MOGAD Diagnostic Criteria (Banwell et al. 2023)). Patients were required to have received at least one dose of rituximab or ocrelizumab between January 2016 and March 2024, with no switch to another disease-modifying therapy or immunotherapy, including a different anti-CD20 agent, at any point since the commencement of therapy up to the end of the outcome-assessment period. This study was approved by the King Saud Medical City Institutional Review Board (approval number: B0AI-03-Oct24-01); informed consent was waived, as the study posed no more than minimal risk to participants.

The primary outcome was to determine the proportion of patients who had LON, defined as an ANC 1.5 Œ 10^9^/L occurring more than 4 weeks after the preceding anti-CD20 infusion, without an alternative explanation, on a complete blood count with differential (CBCD) drawn between April 2023 and April 2024 (a 13-month period). Routine practice at our center included a CBCD blood draw within a 12-week window prior to each infusion; because some patients received rituximab only once yearly, the minimum number of ANC assessments per patient during the study period was one.

Neutropenia severity was graded 1–4 by ANC: Grade 1, 1.0–<1.5 Œ 10^9^/L; Grade 2, 0.5–<1.0 Œ 10^9^/L; Grade 3, 0.1–<0.5 Œ 10^9^/L; Grade 4, <0.1 Œ 10^9^/L (adapted from CTCAE version 6.0).

Episodes were described in terms of their relation to the preceding infusion, symptom status, grade, neutropenia-directed treatment, recovery, re-dosing with anti-CD20, and recurrence. An episode was classified as asymptomatic if the patient had no clinical symptoms attributable to neutropenia (e.g., fever, infection), irrespective of whether treatment (e.g. G-CSF) was administered.

We also investigated demographic and clinical factors, prior disease-modifying therapy, and the specific anti-CD20 agent for their association with neutropenia.

Because of this limited observation window, the analysis was primarily descriptive; inferential statistics presented were exploratory regardless of statistical significance, and were intended to justify more extensive evaluation in future work.

Descriptive statistics were used to summarize data. Continuous variables were summarized as mean ś SD or median (IQR), selected according to normality (Shapiro-Wilk), with the number of missing observations reported separately.

Categorical variables were summarized as the number (n) and percentage (%) of patients in each category; 95% CIs were presented where appropriate.

The proportion of patients with LON was calculated as the number of LON cases identified during the study period, divided by the total number of patients in the cohort, expressed as a percentage; its 95% CI was calculated using the Wilson score method.

Chi-square test or Fisher’s exact test (2Œ2 tables), or the Fisher-Freeman-Halton exact test estimated by Monte Carlo simulation (100,000 replicates) for larger tables with low expected cell counts, and the Welch t-test or Mann-Whitney U test (for continuous variables, according to normality) were used, as appropriate, to compare patients with and without neutropenia.

All analyses were performed in Python 3.12.12, using pandas (2.3.3), numpy (2.4.2), matplotlib (3.10.8), scipy (1.17.1), and statsmodels (0.14.6).

## 3 Results

A total of 229 patients had received at least one dose of rituximab or ocrelizumab on file; 8 were excluded because anti-CD20 therapy was initiated outside the study exposure window (January 2016–March 2024), leaving 221 patients. Of these, 4 were excluded for missing demographic or infusion records, or unverifiable infusion history on chart review, leaving 217 patients with verified demographic and infusion records. Finally, 46 patients had no labs during the study period (7 with no laboratory data on file at all, and 39 with laboratory data on file but no post-treatment blood draw during the study window), leaving 171 patients for analysis (Figure 1). No patient under active follow-up switched to another disease-modifying therapy during the study period; the 46 patients above may include those who discontinued anti-CD20 therapy, switched agents, or transferred care elsewhere without a subsequent documented visit at our center, which could not be independently confirmed from the absence of a qualifying blood draw alone. Rituximab was administered to 141 patients and ocrelizumab to 30 patients.

**Figure 1:**
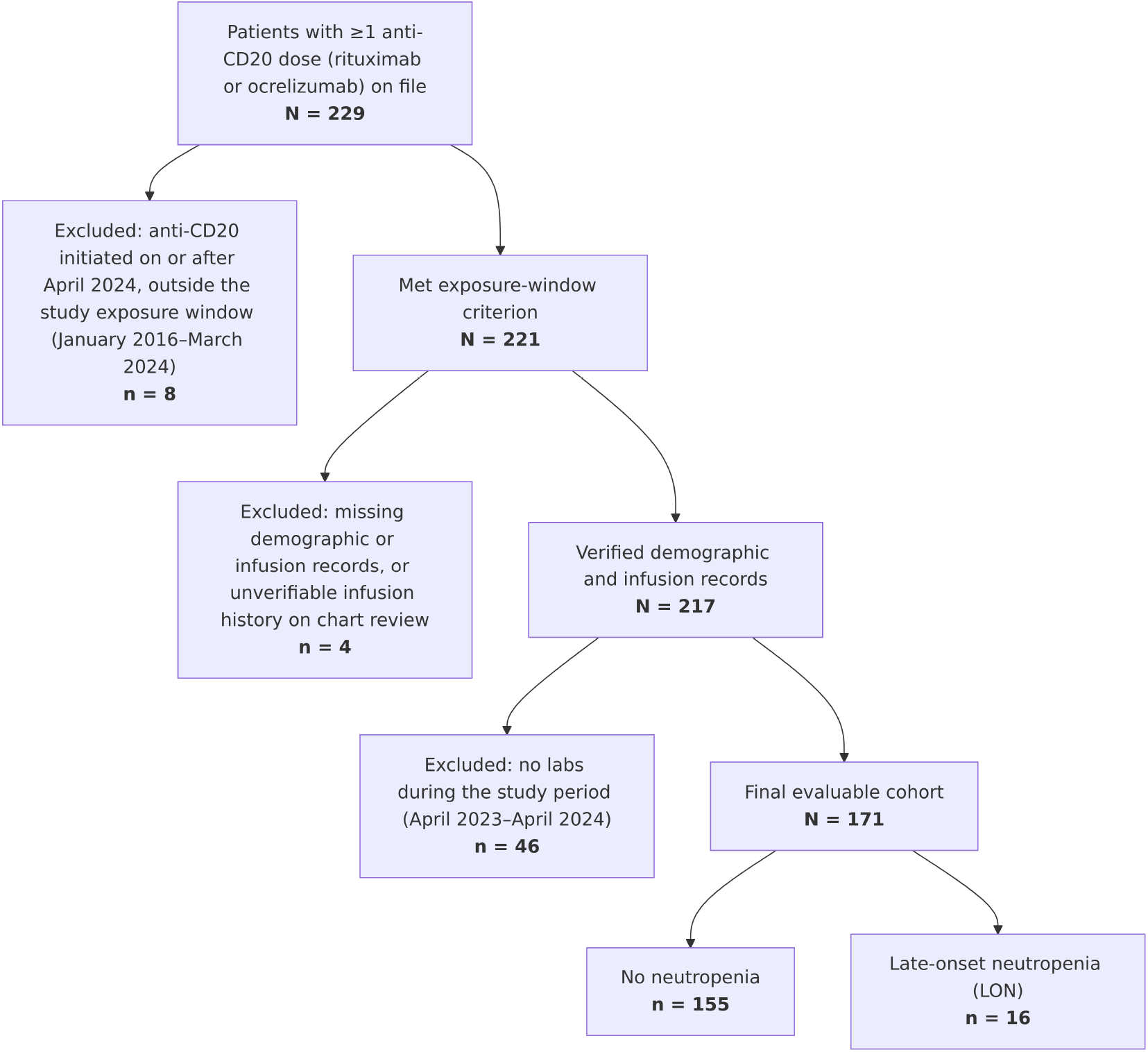
Patient enrollment and exclusion flow diagram.

A total of 385 ANC tests were performed on the final evaluable cohort between April 4, 2023 and April 29, 2024; 319 were post-treatment tests (the remainder were pre-treatment baseline draws). The observation period for the outcome spanned 13 months. Each patient underwent a median of 2.0 (1.0–2.0) tests, with patients who had neutropenia undergoing significantly more tests than those who did not (2.0 (2.0–2.0) vs. 2.0 (1.0–2.0); p = 0.010). A total of 13/319 (4.1%) blood draws happened at an interval of more than 365 days since the last infusion, belonging to 12/171 (7.0%) patients.

Neutropenia was observed in 16/171 (9.4%, 95% CI 5.8–14.7) patients, 12 on rituximab and 4 on ocrelizumab. A detailed description of the baseline characteristics of the cohort is provided in Table 1.

**Table 1:** Baseline characteristics of the cohort.

| Variable | Whole<br>neutropenia<br>cohort | Whole<br>no-neutropenia<br>cohort | p-<br>value | Rituximab<br>(RTX) | Ocrelizumab<br>(OCR) |
| --- | --- | --- | --- | --- | --- |
| N | 16 | 155 |  | 12 | 4 |
| Age at anti-CD20 start, years,<br>Median (IQR) | 29.8 (24.9–35.0) | 31.2 (25.6–36.6) | 0.363 | 28.2<br>(23.6–35.0) | 30.8<br>(29.6–34.0) |
| Disease duration, years,<br>Median (IQR) | 3.3 (0.3–5.3) | 1.4 (0.2–4.2) | 0.379 | 2.2 (0.1–5.3) | 3.4 (2.9–5.1) |
| BMI, kg/m<sup>2</sup>, Median (IQR) | 22.2 (18.8–24.3) | 27.5 (22.7–33.2) | 0.001 | 21.8<br>(18.8–23.2) | 24.0<br>(22.2–24.9) |
| Missing (BMI, kg/m<sup>2</sup>), n (%) | 2 (12.5%) | 43 (27.7%) |  | 2 (16.7%) | 0 |
| Gender |  |  | 1.000 |  |  |
| Female | 12 (75.0%) | 114 (73.5%) |  | 9 (75.0%) | 3 |
| Disease phenotype |  |  | 1.000 |  |  |
| RRMS | 13 (81.2%) | 129 (83.2%) |  | 11 (91.7%) | 2 |
| PPMS | 1 (6.2%) | 10 (6.5%) |  | 0 | 1 |
| Other | 2 (12.5%) | 16 (10.3%) |  | 1 (8.3%) | 1 |
| DMT prior to anti-CD20 |  |  | 0.023 |  |  |
| No therapy | 8 (50.0%) | 72 (46.5%) |  | 7 (58.3%) | 1 |
| Natalizumab | 6 (37.5%) | 22 (14.2%) |  | 3 (25.0%) | 3 |
| Other | 2 (12.5%) | 61 (39.4%) |  | 2 (16.7%) | 0 |

*Continuous variables reported as median (IQR) given non-normal distributions (Shapiro-Wilk); comparisons used the Mann-Whitney U test (continuous) or chi-square/Fisher’s exact test (categorical), comparing the whole neutropenia cohort vs. the whole no-neutropenia cohort. The rituximab (RTX) and ocrelizumab (OCR) columns describe the neutropenia cohort split by anti-CD20 agent (descriptive only, no p-value); percentages are omitted for the ocrelizumab column (n=4) given the small denominator, and raw counts are shown instead. Disease phenotype “Other” (n=18 cohort-wide) comprises secondary progressive MS, NMOSD (seropositive and seronegative), Susac syndrome, pachymeningitis, and CNS vasculitis, combined because each individual category had fewer than 10 patients cohort-wide, to protect patient identifiability. DMT prior to anti-CD20 “Other” (n=63 cohort-wide) comprises fingolimod, betaferon, avonex, teriflunomide, dimethyl fumarate, MMF, oral steroid, azathioprine, rebif, and siponimod, collapsed given their low individual frequency. Rows with zero missing values in every column are omitted. All comparisons are exploratory/hypothesis-generating and not adjusted for multiplicity.*

The median age was 29.8 (24.9–35.0) years, and 12 (75.0%) were female. The majority were diagnosed with relapsing-remitting multiple sclerosis (13, 81.2%). Body mass index (BMI) and DMT prior to anti-CD20 were the two variables that differed significantly between patients who had neutropenia and those who did not: BMI was lower in the neutropenia group (22.2 (18.8–24.3) vs. 27.5 (22.7–33.2); p = 0.001) (Figure 2), and prior natalizumab exposure was more common in the neutropenia group (6/16, 37.5% vs. 22/155, 14.2%; p = 0.023). Among the 28 patients with prior natalizumab exposure, the interval to anti-CD20 initiation did not differ significantly between groups (neutropenia 48.5 days (29.8–74.0) vs. no neutropenia 48.0 days (42.0–91.0); p = 0.416). Among the 6 natalizumab-prior neutropenia cases specifically, the interval from the last natalizumab dose to the first neutropenia episode ranged widely, from 78 to 1568 days (median 1171, IQR 428.5–1403.5); only 1/6 (16.7%) occurred within 6 months of the last natalizumab dose. Exploratory analysis of number of anti-CD20 infusions, cumulative dose, and time on treatment showed no association with the primary outcome (p = 0.728, 0.985, and 0.989; Supplementary Table 1).

**Figure 2:**
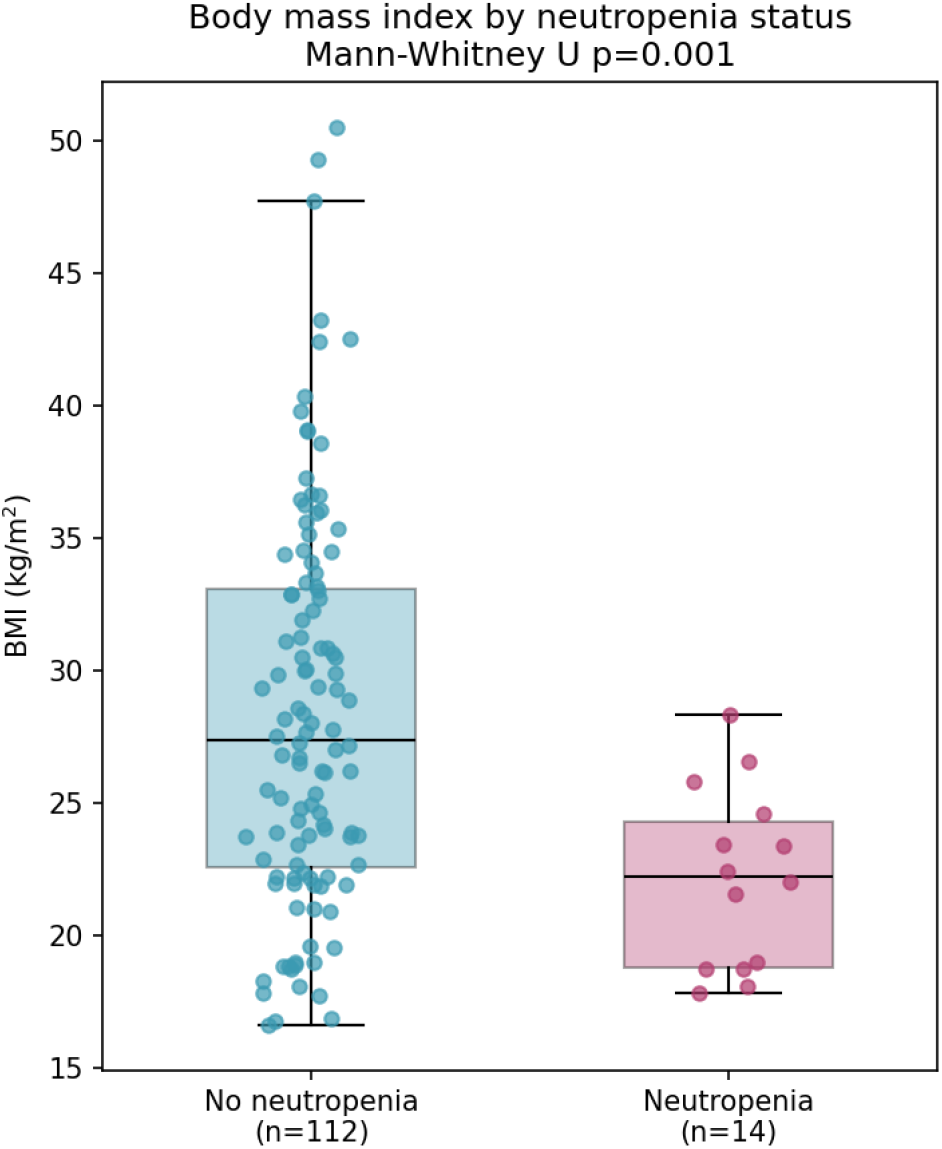
Body mass index (BMI) by neutropenia status. Box plots show median and interquartile range (whiskers to 1.5ŒIQR, outliers not shown); individual patients are overlaid as jittered points. Neutropenia, n=14 with BMI recorded (reddish-purple); no neutropenia, n=112 with BMI recorded (bluish-green). Median BMI 22.2 vs. 27.5 kg/mš (Mann-Whitney U p=0.001).

The proportion of patients with LON in the whole cohort was 9.4% (95% CI 5.8–14.7); for the breakdown by agent, see Table 2.

**Table 2:**
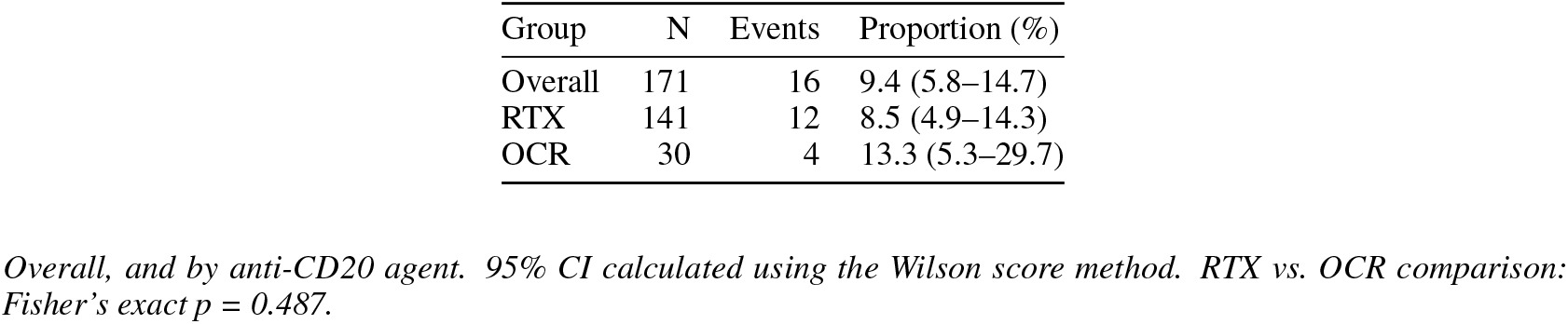
Proportion with late-onset neutropenia (LON).

All these episodes were asymptomatic and did not require any medical intervention. These episodes occurred at a median of 158 days (130–188) since the last anti-CD20 infusion. The majority were Grade 1 (15/16, 93.8%); a single Grade 2 case occurred on ocrelizumab. Documentation of recovery on repeat ANC testing during the study period was available for 8/16 (50.0%) episodes. All but one patient were dosed again after the documentation of neutropenia (15/16, 93.8%) (Table 3). A detailed description of these events is available in Table 4.

**Table 3:** Summary of neutropenia episode characteristics.

| Variable | Overall | Rituximab (RTX) | Ocrelizumab (OCR) |
| --- | --- | --- | --- |
| N episodes | 16 | 12 | 4 |
| Days since preceding infusion, Median (IQR) (n=16) | 157.5 (130.2–188.2) | 151.5 (87.5–171.2) | 261.5 (180.0–350.0) |
| Episode duration (days), Median (IQR) (n=16) | 0.0 (0.0–9.8) | 0.0 (0.0–14.2) | 0.0 (0.0–9.8) |
| Time to recovery (days), Median (IQR) (n=8) | 59.0 (28.5–170.8) | 53.5 (13.5–143.0) | 114.5 (80.8–148.2) |
| Grade |  |  |  |
| Grade 1 | 15 (93.8%) | 12 (100.0%) | 3 |
| Grade 2 | 1 (6.2%) | 0 (0.0%) | 1 |
| Recovered (next ANC $\geq 1.5$) | | | |
| Yes | 8 (50.0%) | 6 (50.0%) | 2 |
| No | 0 (0.0%) | 0 (0.0%) | 0 |
| Unknown (no further labs) | 8 (50.0%) | 6 (50.0%) | 2 |
| Dosed again after event |  |  |  |
| Yes | 15 (93.8%) | 11 (91.7%) | 4 |
| No | 1 (6.2%) | 1 (8.3%) | 0 |
| Repeated neutropenia episodes |  |  |  |
| No | 16 (100.0%) | 12 (100.0%) | 4 |

Overall, and by anti-CD20 agent; percentages are omitted for the ocrelizumab column (n=4) given the small denominator, and raw counts are shown instead. “Unknown (no further labs)” indicates no post-episode ANC test was available within the study period to confirm recovery status — this reflects the fixed observation window, not a failure to follow up; “No” (ANC did not recover to 1.5) did not occur in this cohort but is shown for completeness.

**Table 4:**
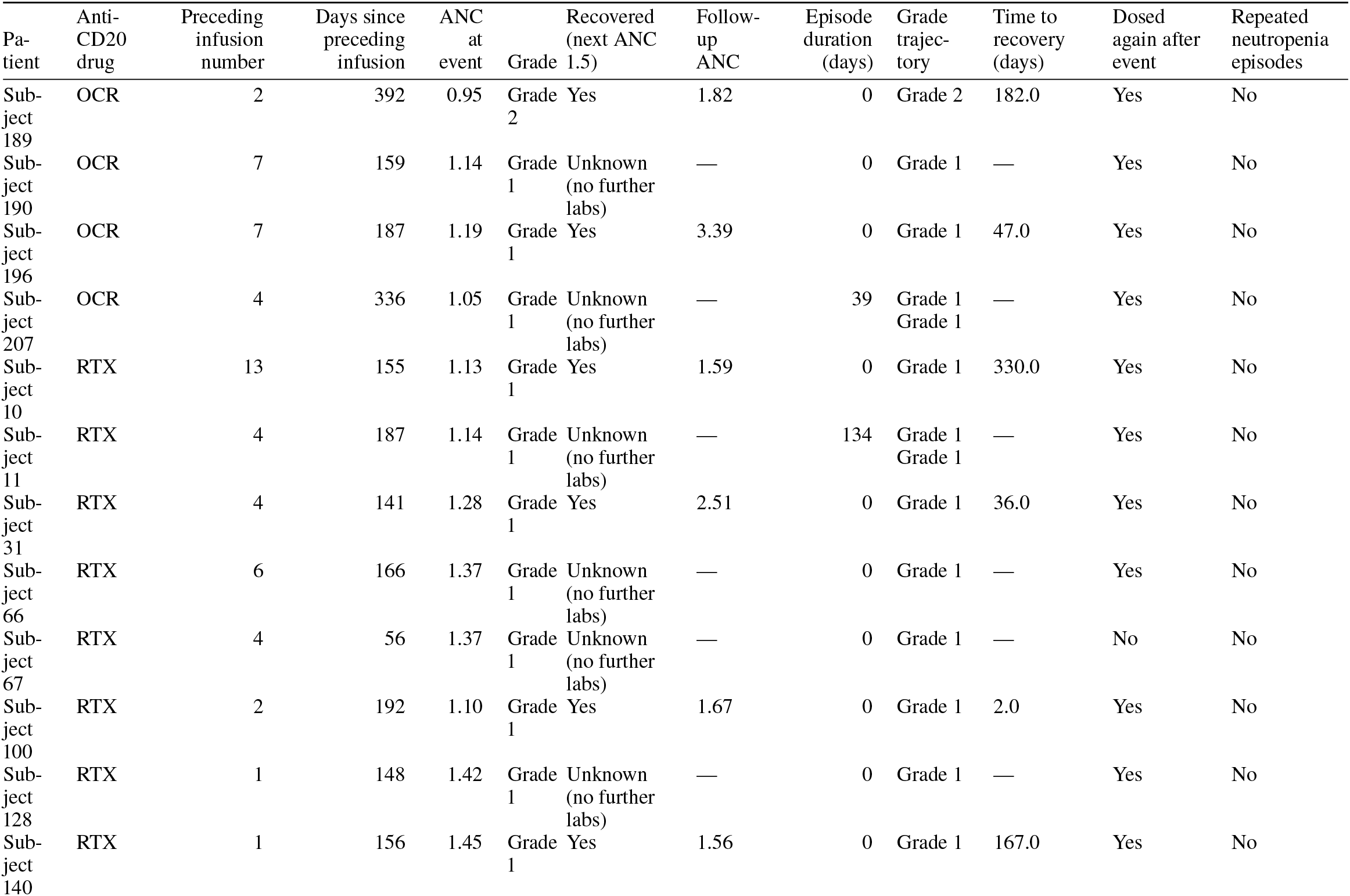
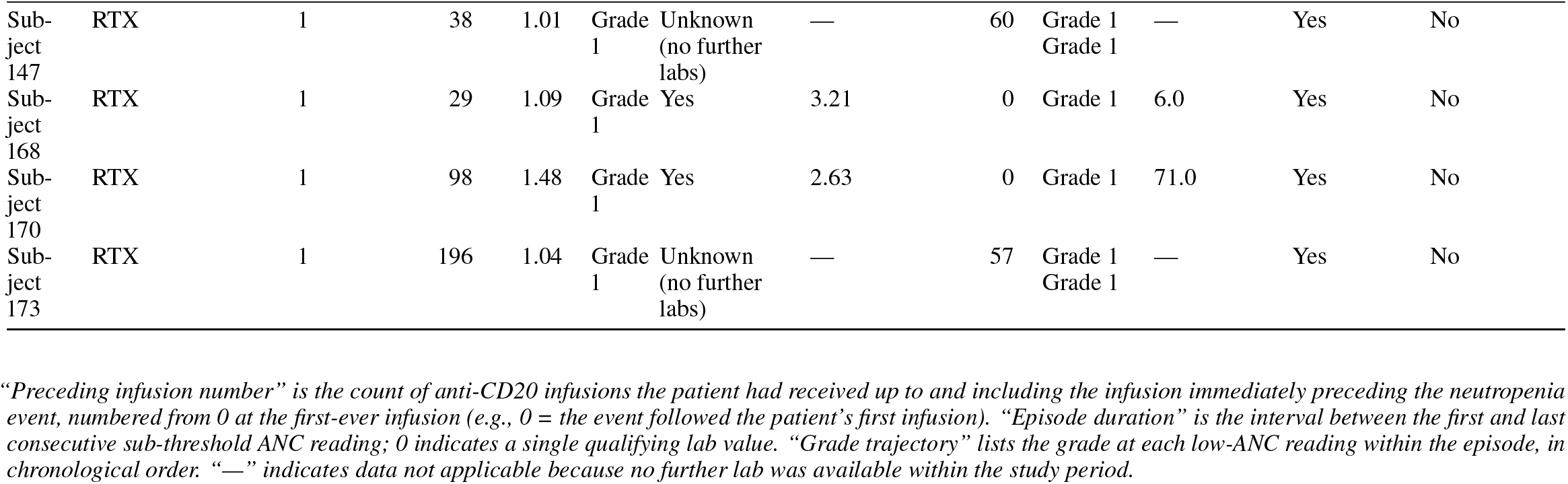
Detailed description of each neutropenia episode (n=16).

## 4 Discussion

Over our 13-month observation period, the proportion of patients with LON in our cohort (9.4%, 95% CI 5.8–14.7) was notably higher than most previously reported cohorts.

Rigal et al. (Rigal et al. 2020) reported a proportion of 2.6% (10/385) over a 12-year retrospective review (2007–2019), with systematic blood testing every 3 months in rituximab-treated patients with CNS autoimmunity. In a subsequent prospective study designed to detect more asymptomatic cases, Rigal et al. (Rigal et al. 2022) performed CBCD every 2 weeks for 6 months from anti-CD20 initiation (rituximab or ocrelizumab) and found a proportion of 1.32% (2/152); however, their ANC threshold (<1.0Œ10^9^/L) excludes Grade 1 cases (1.0–1.5Œ10^9^/L). Waldrop et al. (Waldrop et al. 2025), over nearly 6 years of observation, reported an incidence rate of 0.62 (95% CI 0.45–0.85) per 100 person-years across all B-cell-depleting therapies (rituximab, ocrelizumab, ofatumumab, ublituximab, inebilizumab) for CNS autoimmunity, identifying 37 LON episodes among 1825 patients over the review period. A similarly low proportion, 1.7% (2/117), was reported by (Silva et al. 2026) in ocrelizumab-treated patients, though their observation period was not specified.

In fact, our proportion is not entirely unprecedented: Sartori et al. (Sartori et al. 2025) reported an overall neutropenia proportion of 21.9% (16/73) in their ocrelizumab-treated multiple sclerosis cohort using a broader ANC threshold (<2.0Œ10^9^/L); restricted to ANC <1.5Œ10^9^/L, matching our own case definition, their proportion was 9.6% (7/73) — nearly identical to ours (9.4%).

Population-level differences in baseline neutrophil counts may also contribute to the higher proportion observed here relative to comparator cohorts drawn predominantly from European and North American populations. Benign ethnic neutropenia is common in populations of African, Middle Eastern, and Mediterranean ancestry and is associated with a lower normal range for absolute neutrophil count without any associated immune deficit (Reich et al. 2009). In a large Saudi laboratory cohort, the 2.5th percentile of neutrophil count fell below the standard lower limit of 1.5Œ10^9^/L across all sampled regions, and isolated mild-to-moderate neutropenia was found in 11–23% of routine complete blood counts (Awan et al. 2021). Because our case definition used a fixed absolute threshold rather than each patient’s own baseline ANC, some fraction of the predominantly Grade 1 cases identified here (15/16, 93.8%; ANC 1.0–1.5Œ10^9^/L) may reflect constitutional variation in neutrophil count rather than a drug-induced effect, and this could not be distinguished from true late-onset neutropenia within our study design.

Although the trigger for LON is not established for this treatment class, dosing regimen and route of administration have been proposed as contributing factors (Baker et al. 2024).

BMI was significantly lower in the neutropenia group; each 1-unit increase in BMI was associated with a 17% reduction in the odds of LON in univariable logistic regression (OR 0.83, 95% CI 0.73–0.95, p=0.005). BMI was missing in 45/171 patients (26.3%), disproportionately among patients without LON (Supplementary Table 3); repeating this analysis with missing BMI values multiply imputed left the association materially unchanged (OR 0.84, 95% CI 0.75–0.96, p=0.009, full cohort of 171 patients; Supplementary Table 4), arguing against the missing-data pattern as an explanation for the finding. Waldrop et al. (Waldrop et al. 2025) similarly observed a lower BMI in their neutropenia group, though the difference did not reach statistical significance. Silva et al. (Silva et al. 2026) reported two ocrelizumab-treated patients with LON who had BMIs of 16 kg/mš and 20 kg/mš, consistent with a possible link between lower body weight and LON risk.

The BMI association may also relate to dosing practice: ocrelizumab was dosed on a fixed schedule throughout the study period, while rituximab maintenance dosing for multiple sclerosis shifted between fixed-dose and body-surface-area-based regimens over 2016–2024. Fixed-dose regimens deliver a higher effective per-kilogram dose to lower-BMI patients, offering a plausible but untested pharmacokinetic explanation for the BMI association.

Prior natalizumab exposure was associated with LON in our cohort: 6/16 (37.5%) of neutropenia cases had received natalizumab as their prior DMT, compared to 22/155 (14.2%) of the no-neutropenia group. Expressed as the neutropenia rate within each exposure group, this association was directionally consistent across several reasonable ways of grouping DMT categories and restricting the cohort: natalizumab vs. all other prior DMTs combined (21.4% vs. 7.0%, p=0.028), restricted to multiple sclerosis diagnoses only (21.4% vs. 7.0%, p=0.030), and further excluding DMTs not used for MS (21.4% vs. 7.1%, p=0.032) — all nominally significant (Supplementary Table 2). These restrictions are overlapping subsets of the same 171 patients rather than independent replications and should not be read as separate confirmatory evidence. Against the single most clinically relevant comparator, no prior DMT at all, the contrast was directionally the same but did not reach significance in this small subgroup (natalizumab 21.4% vs. no prior therapy 10.0%, p=0.187), indicating that some of the significance in the aggregated comparison reflects the heterogeneous “other DMT” category running lower rather than natalizumab alone running higher. The association was only lost when DMT-prior categories were left fully disaggregated into their 12 individual agents (p=0.383), which we attribute to power dilution.

Because natalizumab-prior patients in our cohort also ran a lower median BMI (22.2 vs. 27.4 kg/mš cohort-wide), we fit a multivariable logistic regression model including both BMI and natalizumab-prior status to assess whether either association was simply a proxy for the other; both remained significant after mutual adjustment (BMI: OR 0.85, 95% CI 0.74–0.97, p=0.014; natalizumab-prior: OR 4.16, 95% CI 1.17–14.86, p=0.028; n=126, 14/16 LON events with BMI available), suggesting the two are at least partially independent risk markers rather than one fully explaining the other.

We have no plausible mechanistic explanation. Natalizumab is well known to induce hematological changes not seen in normal conditions, including reduced retention of progenitor cells in the bone marrow: Bridel et al. (Bridel et al. 2015) showed that neutrophil progenitor cells are detectable in the peripheral blood of natalizumab-treated patients, persistently elevated over 18 months of treatment. If this mechanism underlies our finding, LON would be expected to cluster within a comparable timeframe after the last natalizumab dose. It did not: among the 6 natalizumab-prior neutropenia cases, the interval from the last natalizumab dose to the first neutropenia episode ranged from 78 to 1568 days (median 1171, IQR 428.5–1403.5), and only 1/6 (16.7%) occurred within 6 months of the last natalizumab dose. This timing rules out a persisting bone-marrow effect from natalizumab itself as the explanation, without calling the natalizumab–LON association itself into question. Switching from natalizumab to anti-CD20 therapy is a common clinical strategy (Zanghi et al. 2021; Bsteh et al. 2025); if this mechanism were responsible, that practice would be expected to have produced a correspondingly higher rate of LON reports following such switches, which has not been reported in the literature — though this absence may equally reflect that case reports of LON rarely stratify by prior DMT, rather than genuine absence of the phenomenon.

Studies with more frequent, protocol-based testing appear to detect a higher proportion of asymptomatic LON, compared to studies relying on symptom-driven or infrequent, irregular testing. In our cohort and in Sartori et al.’s (Sartori et al. 2025) — both retrospective designs with routine, protocol-based blood draws collected within the treating institution — all detected cases were asymptomatic. Rigal et al.’s prospective study (Rigal et al. 2022), which used biweekly CBCD specifically to detect subclinical cases, similarly found both of its cases asymptomatic. By contrast, studies relying on symptom-driven or infrequent testing reported a substantially smaller asymptomatic fraction: Waldrop et al. (Waldrop et al. 2025) (12/37, 32.0%), Silva et al. (Silva et al. 2026) (10/37, 27.0%), and Rigal et al.’s retrospective cohort (Rigal et al. 2020) (4/10, 40.0%). This pattern suggests that testing frequency determines whether LON is captured while still asymptomatic.

As of today, it is not clearly established whether all cases of LON arise from the same mechanism, as several plausible mechanisms exist (Baker et al. 2024). The ability to predict — and therefore mitigate — this risk would be of great clinical value. Better understanding of LON’s natural history and potentially heterogeneous mechanisms could help reduce unnecessary medical intervention.

This study has several limitations. It was conducted at a single center, limiting generalizability. Blood draws were not performed on a standardized schedule but followed routine clinical practice, typically anchored to the next scheduled infusion; this confounds the interval between infusion and neutropenia detection with the interval to the next test, and could not be disentangled from a true biological time course in this cross-sectional design. Related to this, patients who developed LON were tested significantly more often than those who did not (though not over a significantly longer span), raising the possibility of ascertainment bias inflating the apparent proportion relative to a population under uniform surveillance. As a bound on this effect, restricting to only the first qualifying post-treatment test per patient — removing any advantage from repeated testing — yielded a lower but still materially elevated proportion of 6.4% (95% CI 3.6–11.2%; Supplementary Table 5), still exceeding every previously reported estimate cited above (1.32–2.6%), indicating the elevated proportion is not solely attributable to differential testing frequency. No mechanism was directly investigated; the natalizumab and BMI associations discussed above are hypothesis-generating and require confirmation. BMI was calculated at the baseline visit; longitudinal weight change and its relation to outcome were not assessed. Multiple comparisons were performed without correction for multiplicity; all reported associations should be interpreted as exploratory rather than confirmatory. Finally, the cohort included a relatively small number of ocrelizumab-treated patients, limiting the power of by-drug comparisons, and did not include other anti-CD20 agents (e.g., ofatumumab, ublituximab), limiting generalizability beyond rituximab and ocrelizumab.

This study also has several strengths. It provides granular, episode-level detail for each case of LON, including grading, trajectory, recovery, and redosing outcomes. Although our proportion estimate is higher than most prior reports, a nearly identical proportion was independently reported by Sartori et al. (Sartori et al. 2025) once a matched case definition was applied (9.6% vs. our 9.4%). This single concordant comparator does not resolve the disagreement with the majority of cited studies, which report proportions 3-to 7-fold lower, and could equally reflect methodological features shared between the two cohorts (both used routine, protocol-based institutional testing) rather than confirming a higher true population proportion; we present it as one data point consistent with our finding, not as its confirmation. The BMI and prior natalizumab associations identified here are hypothesis-generating and may prompt further investigation of anti-CD20 dosing strategy and DMT-switching practices as risk factors. This cohort also draws from a Middle

Eastern population, adding representation of an understudied population to the existing LON literature. Finally, we applied a single, standard case definition (CTCAE v6-based ANC threshold) consistently throughout, unlike several cited comparators that mix inconsistent thresholds and grading conventions.

## 5 Conclusion

Our cohort of rituximab and ocrelizumab-treated CNS autoimmunity patients shows a high proportion of patients with LON (16/171, 9.4%) compared to most previously reported cohorts (1.32–2.6%; one study using a matched case definition reported a nearly identical proportion of 9.6%). All cases were asymptomatic, suggesting asymptomatic LON may be more common than once thought. Lower BMI and prior natalizumab exposure emerged as potential risk factors, warranting further investigation. A larger, prospective study with a standardized blood-testing protocol can establish the true frequency, identify individuals at risk, and determine contributing factors.

## 7 Declarations

### 7.1 CRediT authorship contribution statement

**Ahmed H. Althobaiti:** Conceptualization, Methodology, Software, Formal analysis, Investigation, Data curation, Visualization, Writing – original draft, Writing – review & editing, Supervision, Project administration.

**Nora A. Abanmi:** Data curation, Investigation, Methodology, Resources, Validation, Writing – review & editing.

### 7.2 Declaration of generative AI use

During the preparation of this work, the author(s) used a generative AI tool (Claude, Anthropic) at various stages of manuscript preparation to assist with brainstorming and idea generation, drafting, language, and editing. All AI-generated content was iteratively reviewed, verified against source data, and edited by the author(s) to ensure accuracy. The author(s) take full responsibility for the content of the published article.

### 7.3 Ethics Statement

This study was approved by the King Saud Medical City Institutional Review Board (approval number: B0AI-03-Oct24-01). Patient informed consent was not required, as data were collected exclusively from routine clinical records with no patient contact initiated for research purposes.

### 7.6 Data Availability Statement

The dataset supporting the conclusions of this article is available from the corresponding author upon reasonable request and subject to institutional data governance approval.

