## supplementary table for "Late-Onset Neutropenia in a Single-Center, Retrospective Cohort of Central Nervous System Autoimmunity Patients Treated with Anti-CD20"

### Supplementary Materials

Supplementary Table 1: Exposure to anti-CD20 therapy (number of infusions, cumulative dose, and time on treatment), up to the neutropenia event or last available follow-up.

| Variable | Whole neutropenia cohort | Whole no-neutropenia cohort | p-value | Rituximab (RTX) | Ocrelizumab (OCR) |
|---|---|---|---|---|---|
| N | 16 | 155 | | 12 | 4 |
| Number of infusions, Median (IQR) | 3.5 (2.0–5.5) | 4.0 (2.0–6.0) | 0.728 | 2.5 (2.0–5.0) | 6.5 (4.5–8.0) |
| Cumulative anti-CD20 dose, mg, Median (IQR) | 2000.0 (1200.0–4200.0) | 2100.0 (1300.0–3800.0) | 0.985 | 1525.0 (1200.0–3725.0) | 3300.0 (2100.0–4200.0) |
| Time on anti-CD20 before event/last follow-up, months, Median (IQR) | 18.4 (5.5–32.5) | 19.1 (6.1–30.2) | 0.989 | 10.2 (4.9–20.5) | 38.5 (27.8–47.2) |

*Whole neutropenia cohort vs. whole no-neutropenia cohort comparisons used the Mann-Whitney U test. The rituximab (RTX) and ocrelizumab (OCR) columns describe the neutropenia cohort split by anti-CD20 agent (descriptive only, no p-value). Exploratory analysis; not adjusted for multiplicity.*

Supplementary Table 2: Sensitivity analyses for the association between prior natalizumab exposure and LON, across alternative ways of grouping DMT categories and restricting the cohort.

| Restriction | N | Natalizumab, n/N (%) | Other DMTs, n/N (%) | p-value |
|---|---|---|---|---|
| Primary analysis (Table 1 3-category omnibus test): No therapy / Natalizumab / Other, whole cohort | 171 | 6/28 (21.4%) | 10/143 (7.0%) | 0.023 |
| Natalizumab vs. all other DMTs combined (binary), whole cohort | 171 | 6/28 (21.4%) | 10/143 (7.0%) | 0.028 |
| Full ungrouped DMT categories (12 individual agents), whole cohort | 171 | 6/28 (21.4%) | — | 0.383 |

| Restriction | N | Natalizumab, n/N (%) | Other DMTs, n/N (%) | p-value |
|---|---|---|---|---|
| Natalizumab vs. all other DMTs combined (binary), MS diagnoses only | 157 | 6/28 (21.4%) | 9/129 (7.0%) | 0.030 |
| Natalizumab vs. all other DMTs combined (binary), MS diagnoses only, excluding DMTs not used for MS (MMF, oral steroid, azathioprine) | 155 | 6/28 (21.4%) | 9/127 (7.1%) | 0.032 |

*"MS diagnoses only" restricts to relapsing-remitting, secondary progressive, and primary progressive multiple sclerosis, excluding NMOSD, MOGAD, autoimmune encephalitis, Susac syndrome, pachymeningitis, and CNS vasculitis. All comparisons other than the "full ungrouped" row and the primary Table 1 row use a consistent binary grouping (natalizumab vs. every other prior DMT combined, including "no therapy") so that restrictions are directly comparable to one another. Fisher's exact test (2×2 tables) or the Fisher-Freeman-Halton exact test estimated by Monte Carlo simulation (100,000 replicates, seed=42) for larger tables with low expected cell counts, as in Methods. Exploratory/hypothesis-generating; not adjusted for multiplicity.*

Supplementary Table 3: Comparison of patients with missing versus observed BMI, to characterize whether BMI missingness appears related to other measured characteristics.

| Variable | Missing BMI (n=45) | Observed BMI (n=126) | p-value |
|---|---|---|---|
| LON, n/N (%) | 2/45 (4.4%) | 14/126 (11.1%) | 0.243 |
| Age, years, Median (IQR) | 32.2 (26.3–35.9) | 30.3 (25.1–36.1) | 0.504 |
| Disease duration, years, Median (IQR) | 1.4 (0.2–4.6) | 1.5 (0.2–4.2) | 0.554 |
| Female, n (%) | 33 (73.3%) | 93 (73.8%) | 1.000 |
| Ocrelizumab, n (%) | 6 (13.3%) | 24 (19.0%) | 0.496 |
| DMT prior to anti-CD20 | | | 0.389 |
| No therapy | 18 (40.0%) | 62 (49.2%) | |
| Natalizumab | 10 (22.2%) | 18 (14.3%) | |
| Other | 17 (37.8%) | 46 (36.5%) | |
| Disease phenotype | | | 0.633 |
| RRMS | 38 (84.4%) | 103 (81.7%) | |
| PPMS | 2 (4.4%) | 11 (8.7%) | |
| Other | 5 (11.1%) | 12 (9.5%) | |

*Comparisons used Mann-Whitney U (continuous) or Fisher's exact/chi-square test (categorical). BMI was missing in 45/171 patients (26.3%): 2/16 (12.5%) in the LON group and 43/155*

*(27.7%) in the no-LON group. None of the measured covariates differed significantly between patients with missing versus observed BMI, including LON status itself, providing some reassurance against strongly informative missingness, though this cannot rule out missingness related to BMI itself (missing not at random).*

Supplementary Table 4: Sensitivity analysis for the BMI–LON association under multiple imputation of missing BMI values, compared with the complete-case analysis reported in the main text.

| Analysis | N | LON events | OR | 95% CI | p-value |
|---|---|---|---|---|---|
| Complete-case (as reported in main text) | 126 | 14 | 0.83 | 0.73–0.95 | 0.005 |
| Multiple imputation (50 imputations) | 171 | 16 | 0.84 | 0.75–0.96 | 0.009 |

*Univariable logistic regression of LON status on BMI. Missing BMI values were multiply imputed (50 imputations, chained-equations/MICE, statsmodels 0.14.6) using an imputation model that included LON status, age, disease duration, gender, anti-CD20 agent, prior DMT category, and disease phenotype as predictors; estimates were pooled across imputations using Rubin's rules. Fraction of missing information for the BMI coefficient was 0.17, indicating the imputation uncertainty contributed modestly to the overall estimate. The multiple-imputation estimate is materially unchanged from the complete-case estimate, indicating the BMI–LON association is not an artifact of the missing-data pattern. Exploratory analysis; not adjusted for multiplicity.*

Supplementary Table 5: Sensitivity analysis for the overall proportion with LON, restricting to only the first qualifying post-treatment test per patient, to bound the effect of differential testing frequency (ascertainment bias) on the primary estimate.

| Analysis | N | Events | Proportion (%) |
|---|---|---|---|
| As reported (any qualifying post-treatment test) | 171 | 16 | 9.4 (5.8–14.7) |
| First qualifying post-treatment test only | 171 | 11 | 6.4 (3.6–11.2) |

*95% CI calculated using the Wilson score method. The first-test-only analysis simulates a single-snapshot design with no advantage from repeated testing; the five cases lost under this restriction (Subjects 66, 67, 100, 170, 190 in the main-text episode-detail table) were only identified on a second or later qualifying test. Both estimates exceed every previously reported comparator cited in the Discussion (1.32–2.6%). Exploratory analysis; not adjusted for multiplicity.*
